# Correlates of non-dipping blood pressure in persons with and without hypertension

**DOI:** 10.1101/2024.02.25.24303325

**Authors:** Situmbeko Liweleya, Benson M. Hamooya, Obrain M. L. Mungalu, Sharon D. Zimba, Sepiso K. Masenga

**Author notes:** **Address for Correspondence:** Situmbeko Liweleya Mulungushi University School of Medicine and Health Sciences Livingstone, Zambia. **Emails:** Situmbeko Liweleya – Benson M. Hamooya – Obrain M. L. Mungalu – Sharon D. Zimba – Sepiso K. Masenga.

## Abstract

**Background:** Blood pressure (BP) is known to follow a circadian rhythm with 10% to 15% lower values during the night than daytime. Non-dipping BP refers to the absence of BP dipping and has been associated with the development of target organ damage. The general goal of this study was to determine the correlates of non-dipping BP in persons with and without hypertension.

**Methods:** This was a cross-sectional study that recruited 98 participants at Chikankata Mission General Hospital. The outcome variable of the study was non-dipping BP, with sociodemographic and clinical explanatory variables. We used SPSS version 22 to describe and make inferences.

**Results:** The median (interquartile range (IQR)) age of participants was 42 years (34.7-52) and 54.1% (53/98) had hypertension while 45.9% (45/98) were normotensive. The proportion of females was slightly higher (59.2%, n=58) than that of males (40.2%, n=40), this being similar in hypertensives but equal in normotensives. The median (IQR) age of hypertensives was higher compared to the normotensives, 46 (40-56) vs. 35 (25-41) years.

The prevalence of non-dipping BP was 38.8% overall and higher among those with hypertension (54.7%) compared to the normotensive group (20%). The factors associated with non-dipping BP in the multivariate analysis were age (adjusted odds ratios (AOR) of 1.15; 95% CI: 1.05 - 1.25), spot urine sodium (AOR of 1.16; 95% CI: 0.99 - 1.36), daytime systolic blood pressure (SBP) load (AOR of 1.28; 95% CI: 1.06 - 1.55), daytime diastolic blood pressure (DBP) load (AOR of 0.77; 95% CI: 0.65 - 0.92), and nighttime DBP load (AOR of 1.10; 95% CI: 1.02, 1.18). However, this was abrogated by hypertension status albeit among normotensives only age remained significantly associated with non-dipping BP, none of the factors remained significantly associated with non-dipping BP among persons with hypertension.

**Conclusion:** The prevalence of non-dipping BP was high, among hypertensives. This provides insights into the intricate links between BP patterns, sociodemographic and clinical characteristics but further underscores the need for mechanistic researches to further advance the understanding of mechanisms of associated characteristics.

## Introduction

Blood pressure, a crucial indicator of cardiovascular health, is known to undergo predictable variations over a 24-hour cycle, showing higher values during the day and lower values at night, a phenomenon commonly referred to as dipping [1]. Non-dipping blood pressure poses a threat to cardiovascular health, increasing the likelihood of developing conditions such as coronary artery disease, cardiovascular diseases, heart failure, stroke, and mortality [2–4].

Normotensive and hypertensive individuals present with a variation in diurnal rhythm of sodium excretion [5]. The excretion of sodium approaches maximum capacity during daytime and reduces during sleep. Blood pressure has for long been linked as being the primary determinant of nocturnal excretion of sodium [6]. Fukuda *et al.* hypothesized that non-dipping patterns of nocturnal blood pressure were a consequence of impaired capacity to excrete sodium during the night [7]. As such, leading to the demand to maintain a 24-hour sodium balance which in turn is compensated with an increase of blood pressure at night that promotes sodium excretion[7–9]. This impaired capacity to excrete sodium is postulated to be a result of reduced renal function,a characteristic observation in candidates with a low glomerular filtration rate [6]. This hypothesis linking dipping patterns of blood pressure and sodium excretion is fundamentally supported by several studies in selected salt-sensitive and salt resistant patients [10].

Rhythmic blood pressure abnormalities could conceptually arise from a failure in sodium reabsorption that may occur at an inappropriate time of the day [11]. Another possibility may be found in a phenomenon called failure of the reactive system, where a mismatch for sodium ingested, physiological demand and failure in the generation of appropriate reaction. This conceptual framework is then used to explore renal/ sodium-centric mechanism describing the underlying basis of non-dipping blood pressure [12].

Increased nocturnal sodium excretion has a significant association with non-dipping blood pressure [9]. This phenomenon is poorly understood among hypertensive and non-hypertensive black African population. Research studies around the world have consistently reported a prevalence of non-dipping blood pressure in about 74% to 82% of patients with renal impairment [13,14]. Non-dippers tend to exhibit more progressive morphological and functional renal changes compared to dippers [15]. Farmers *et al*. noted an increasing proportion of non-dipping blood pressure corresponding to rising plasma creatinine levels in patients suffering from chronic renal insufficiency [16]. Similarly, Lurbe *et al.* reported a higher incidence of microalbuminuria development in non-dippers with type 1 diabetes mellitus, though this was not observed in those with normal albumin excretion [17]. An attenuation of the typical nighttime dipping, resulting in a non-dipping pattern, has been firmly linked with a heightened risk of hypertension [18].

The main goal of this study was to determine the burden and correlates of nocturnal non-dipping blood pressure in persons with and without hypertension.

## Methods

### Study design and setting

This was a cross-sectional study that was conducted at Chikankata Mission General Hospital, between February to June of 2022, among normotensive and hypertensive individuals. Chikankata Mission General Hospital is a Level 2 Hospital that offers its services to a population of about 98,671 people, operating for a population density of 36.91/km^2^ and 2,673 km^2^ coverage.

### Selection of participants and sampling methods

Participants were selected using convenient sampling during their normal routine checkups in the hospital hypertension clinic population. A control group of normotensive participants were recruited from the general population.

### Eligibility criteria

The study included both normotensive and hypertensive adults aged ≥18 years. Hypertension was defined as systolic blood pressure/diastolic blood pressure (SBP/DBP) of ≥ 140/90 mmHg on more than 2 consecutive occasions or history of antihypertensive medication use. We excluded clients that did not consent, had diabetes and abnormal kidney function and those that were known diabetics. Clients who did not provide consent for participation in the study, as ethical guidelines mandated the necessity of informed consent. Furthermore, individuals with diabetes and abnormal kidney function were excluded from the study due to the potential confounding effects of these conditions on the variables under investigation.

### Sample size estimation

We were planning a study with 42 hypertensives and 48 normotensive individuals to serve as controls. We assumed the maximum probability of non-dipping blood pressure among persons with hypertension to be 50%. If the true probability of non-dipping blood pressure among the controls was 20%, we would be able to reject the null hypothesis that the probability rates for cases and controls are equal with probability (power) 0.858. The Type I error probability associated with this test of this null hypothesis is 0.05. We used an uncorrected chi-squared statistic to evaluate this null hypothesis. In order to account for 10% attrition, we added 4 participants to each group. We therefore recruited 46 persons with hypertension and 52 normotensives.

We used PS Power and Sample Size Calculations found at http://biostat.mc.vanderbilt.edu/PowerSampleSize to conduct power calculation.

### Research Variables

The primary variable of the study was non-dipping blood pressure, which was defined as diminished decrease in blood pressure during nighttime by 10% to 15% [3]. Explanatory Variables included sex, age, hypertension status, smoking, alcohol intake, physical exercises, quality of sleep, BMI, waist circumference, Day SBP load value, Day DBP load value, Night SBP load value, Night DBP load value, spot urine sodium and urine sodium excretion.

SBP and DBP load values refer to the percentage of blood pressure readings above the threshold (typically 140/90 mm Hg for adults). Thus, higher SBP and DBP load values suggest poorer blood pressure control throughout the day and night. We used an ambulatory blood pressure monitor (ABPM50, China) to determine blood pressure loads. Patient preparation was underscored as crucial for accurate ABPM. Patients were guided to stay off caffeine, alcohol, and smoking, dress in comfortable attire, and abstain from physical activities during the testing period. ABPM readings were taken during the patient’s typical awake and sleeping hours, spanning 24 hours. The ABPM device used was a CONTEC (ABPM50, China). The device was to be attached to the non-dominant arm with the cuff securely fitted around the upper arm [19].

To assess sleep quality, the Pittsburgh Sleep Quality Index (PSQI) was used. The PSQI asked questions about sleep disturbances, latency, duration, and other factors to provide a comprehensive score of sleep quality. To assess physical activity, we collected information on the time allocated to different activities, offering a comprehensive measure of the overall engagement duration in physical exercise.

### Antihypertensives

During data collection, the patients diagnosed with hypertension were taking antihypertensive medications, such as a calcium channel blocker (amlodipine or nifedipine), an angiotensin-converting enzyme inhibitor (ACE) (enalapril or losartan), or a diuretic (furosemide or moduretic).

### Spot Urine Sodium Estimation

A Spot urine sample was obtained from participants, collected in a sterile container. To estimate spot urine sodium concentration, we measured sodium concentration using an ion-selective electrode (ISE) CYANSmart CY009 chemistry analyzer and used the INTERSALT formula to estimate spot sodium excreted as shown below:

*Estimated Spot-hour urine sodium excretion (mmol/day) = 14.4 x sodium concentration (mmol/L) x urine volume (L/day)*.

### Data sources/ measurement

We used interviewer structured questionnaires to collect data. These were translated into the local language (Tonga) for participants that could not speak English. Data was deidentified and stored on the principal investigator’s laptop.

### Data Analysis

We used SPSS version 22 statistical software for data analysis. We described continuous variables using median (Interquartile range (IQR)). For categorical variables, we utilized the Chi-square test or Fisher’s exact test where appropriate when comparing with the outcome variable. To determine the correlates of non-dipping blood pressure, we used multivariate logistic regression. P values less than 0.05 were considered significant and are shown in bold.

We have used the Strengthening the reporting of observational studies in epidemiology (STROBE) to guide our reporting, Supplementary file 1.

## Results

### General characteristics of participants

The overall median age of participants was 42 years (IQR 34.7-52.2) with more females (59.2%) than males (40.8%). The majority of participants were hypertensive (54.1%) and non-smokers (91.8%). The prevalence of non-dipping blood pressure was 38.8% (95% CI 29.1 – 49.1). Most of the participants did not take alcohol, did not exercise, had good quality of sleep and had dipping blood pressure. Median BMI, urine sodium, waist circumference, day SBP/DBP and night SBP/DBP are shown in Table 1.

**Table 1.** Study characteristics

| **VARIABLE** | **FREQUENCY/**<br>**MEADIAN** | **PERCENTAGE/ IQR** |
| --- | --- | --- |
| **Age, years (Median/ IQR)** | 42 | 34.7-52.2 |
| **Sex** |  |  |
| *Female* | 58 | 59.2 |
| *Male* | 40 | 40.8 |
| **Hypertension** |  |  |
| *Normotensive* | 45 | 45.9 |
| *Hypertensive* | 53 | 54.1 |
| **Smoking** |  |  |
| *Non-Smoker* | 90 | 91.8 |
| *Smoker* | 8 | 8.2 |
| **Alcohol Intake** |  |  |
| *No* | 84 | 85.7 |
| *Yes* | 14 | 14.3 |
| **Physical Exercise** |  |  |
| *No* | 79 | 80.6 |
| *Yes* | 19 | 19.4 |
| **Quality of Sleep** |  |  |
| *Poor* | 14 | 14.3 |
| *Good* | 84 | 85.7 |
| **Non-Dipping BP** |  |  |
| *Dipping* | 60 | 61.2 |
| *Non-Dipping* | 38 | 38.8 |
| **BMI, $kg/m^2$** | 30.1 | 26.8-32.3 |
| **Spot Urine Sodium, $mmol/l$** | 50.6 | 46.3-53.9 |
| **Waist Circumference, $cm$** | 34.5 | 33.2-35.6 |
| **Urine Sodium Excretion, $mmol/day$** | 143.4 | 133.0-154.6 |
| **Day SBP-Load Value, %** | 7.5 | 5.3-17.7 |
| **Day DBP-Load Value, %** | 9.6 | 6.1-20.3 |
| **Night SBP-Load Value, %** | 4.2 | 2.0-19.4 |
| **Night DBP-Load Value, %** | 5.4 | 2.0-39.6 |

BP; Blood Pressure, SBP; Systolic BP, DBP; Diastolic BP, BMI; Body Mass Index

### Study characteristics showing the burden of nocturnal non-dipping BP by Hypertension Status

In this study, participants with hypertension were older than normotensives with a median age of 46 years (IQR 40-56) and 35 years (IQR 25-41) respectively, P<0.001. Non-dipping BP was significantly associated with hypertension (p-value <0.001). There were more hypertensive females than males. Sex, smoking, alcohol intake, physical exercise and quality of sleep were not associated with hypertension. Participants with hypertension had higher BMI, day SBP/DBP and night SBP/DBP than normotensives, Table 2.

**Table 2:** Study characteristics compared by Hypertension Status

| Variables | Hypertension Status (Median/IQR) (FREQ/%) |  | P-value |
| --- | --- | --- | --- |
|  | Normotensive | Hypertensive |  |
| **Age, years (Median/ IQR)** | 35 (25,41) | 46 (40,56) | **<0.001** |
| **Sex** |  |  | 0.05 |
| *Female* | 22 (48.9) | 36 (67.9) |  |
| *Male* | 23 (51.1) | 17 (32.1) |  |
| **Smoking** |  |  | 0.13 |
| *Non-Smoker* | 39 (86.7) | 51 (96.2) |  |
| *Smoker* | 6 (13.3) | 2 (3.8) |  |
| **Alcohol Intake** |  |  | 0.24 |
| *No* | 41 (91.1) | 43 (81.1) |  |
| *Yes* | 4 (8.9) | 10 (18.9) |  |
| **Physical Exercise** |  |  | 0.88 |
| *No* | 36 (80.0) | 43 (81.1) |  |
| *Yes* | 9 (20.0) | 10 (18.9) |  |
| **Quality of Sleep** |  |  | 0.74 |
| *Poor* | 7 (15.6) | 7 (13.2) |  |
| *Good* | 38 (84.4) | 46 (86.8) |  |
| **Non-Dipping BP** |  |  | **<0.001** |
| *Dipping* | 36 (80.0) | 24 (45.3) |  |
| *Non-Dipping* | 9 (20.0) | 29 (54.7) |  |
| **BMI, kg/m<sup>2</sup>** | 29.0 (24.6,31.3) | 31.1 (29.0,32.4) | **0.001** |
| **Spot Urine Sodium, mmol/l** | 50.0 (46.5,52.7) | 51.0 (46.3,61.0) | 0.19 |
| **Waist Circumference, cm** | 34.0 (32.4,35.4) | 34.8 (33.4,35.8) | 0.08 |
| **Urine Sodium Excretion, mmol/day** | 145.7 (133.9,152.8) | 140.6 (132.6,173.5) | 0.84 |
| **Day SBP-Load Level, %** | 6.9 (3.8,10.5) | 10.1 (5.7,30.9) | **0.003** |
| **Day DBP-Load Level, %** | 7.6 (5.2,13.5) | 9.9 (8.4,30.4) | **0.007** |
| **Night SBP-Load Level, %** | 3.2 (1.6,8.0) | 7.4 (2.4,58.5) | **0.002** |
| **Night DBP-Load Level, %** | 3.1 (1.3,9.8) | 9.7 (2.3,69.6) | **0.001** |

BP; Blood Pressure, SBP; Systolic BP, DBP; Diastolic BP, BMI; Body Mass Index

**Table 2.** Study characteristics comparing dipper and non-dippers

| VARIABLES | NON-DIPPING BP (MEDIAN/IQR) |  | P-VALUE |
| --- | --- | --- | --- |
|  | DIPPING | NON-DIPPING |  |
| **Age, years** | 38 (32,53.3) | 53 (46,61.2) | **<0.001** |
| **Sex** |  |  | 0.13 |
| Female | 32 (53.3) | 26 (64.4) |  |
| Male | 28 (46.7) | 12 (31.6) |  |
| **Smoking** |  |  | 1.00 |
| Non-Smoker | 55 (91.7) | 35 (92.1) |  |
| Smoker | 5 (8.3) | 3 (7.9) |  |
| **Alcohol Intake** |  |  | 0.80 |
| No | 51 (85.0) | 33 (86.8) |  |
| Yes | 9 (15.0) | 5 (13.2) |  |
| **Physical Exercise** |  |  | 0.21 |
| No | 46 (76.7) | 33 (86.8) |  |
| Yes | 14 (23.3) | 5 (13.2) |  |
| **Quality of Sleep** |  |  | 0.55 |
| Poor | 10 (16.7) | 4 (10.5) |  |
| Good | 50 (83.3) | 34 (89.5) |  |
| **Hypertension Status** |  |  | **<0.001** |
| Normotensive | 36 (60.0) | 9 (23.7) |  |
| Hypertensive | 24 (40.0) | 29 (76.3) |  |
| **BMI, kg/m<sup>2</sup>** | 29.7 (25.3,32.4) | 30.6 (28.0,31.7) | 0.62 |
| **Spot Urine Sodium, mmol/l** | 49.1 (46.0,52.3) | 52.3 (47.9,65.3) | **0.006** |
| **Waist Circumference, cm** | 34.3 (32.7,35.6) | 34.8 (33.4,36.1) | 0.10 |
| **Urine Sodium Excretion, mmol/day** | 140.6 (130.7,150.4) | 149.2 (136.9,200.8) | **0.009** |
| **Day SBP-Load Level, %** | 6.1 (3.8,9.2) | 15.9 (6.6,47.4) | **<0.001** |
| **Day DBP-Load Level, %** | 7.8 (5.1,10.7) | 18.3 (8.9,48.1) | **<0.001** |
| **Night SBP-Load Level, %** | 2.6 (1.7,5.6) | 18.8 (7.3,92.3) | **<0.001** |
| **Night DBP-Load Level, %** | 2.3 (1.6,6.7) | 49.6 (9.4,87.3) | **<0.001** |

BP; Blood Pressure, SBP; Systolic BP, DBP; Diastolic BP, BMI; Body Mass Index

### Study characteristics showing the burden of nocturnal non-dipping BP

Participants with non-dipping BP were older than those with dipping BP with a median age of 53 years (IQR 46-61.2) and 38 years (IQR 32-53.3) respectively, p<0.001. The prevalence of non-dipping BP was higher in hypertensives compared to normotensives, 76.3% vs 23.7%. Age, hypertension status, urine sodium, urine sodium excretion, day/night SBP and day/night DBP were all significantly associated with non-dipping BP. However, sex, smoking, alcohol intake, physical exercise and quality of sleep were not associated with non-dipping BP. Participants with non-dipping BP had higher urine sodium, urine sodium excretion, day SBP, night SBP and night DBP than those with dipping BP and this association was statistically significant, p<0.05, see Table 3.

**Table 3.** Factors associated with non-dipping BP in the study population

| Variables | OR (95%CI) | P value | AOR (95%CI) | P value |
| --- | --- | --- | --- | --- |
| **Age, years** | 1.16 (1.09,1.24) | **<0.001** | 1.15 (1.05,1.25) | **0.002** |
| **Sex** |  | 0.14 |  | 0.67 |
| *Females* | 0.52 (0.22,1.23) |  | 1.48 (0.24,9.12) |  |
| **Hypertension Status** |  | **<0.001** |  | 0.30 |
| *Hypertensive* | 4.83 (1.94-11.9) |  | 0.36 (0.05,2.48) |  |
| **Spot Urine Sodium, mmol/l** | 1.11 (1.04,1.18) | **<0.001** | 1.16 (0.99,1.36) | 0.05 |
| **Waist Circumference, cm** | 1.21 (1.01,1.45) | **0.04** | 1.49 (0.96,2.31) | 0.07 |
| **Urine Sodium Excretion, mmol/day** | 1.03 (1.01,1.04) | **<0.001** | 1.03 (0.98,1.08) | 0.13 |
| **Day SBP Load Level, %** | 1.07 (1.03,1.12) | **<0.001** | 1.28 (1.06,1.55) | **0.009** |
| **Day DBP Load Level, %** | 1.04 (1.01,1.07) | **0.002** | 0.77 (0.65,0.92) | **0.004** |
| **Night SBP Load Level, %** | 1.04 (1.02,1.07) | **<0.001** | 0.94 (0.86,1.04) | 0.26 |
| **Night DBP Load Level, %** | 1.04 (1.02,1.05) | **<0.001** | 1.10 (1.02,1.18) | **0.002** |

BP; Blood Pressure, SBP; Systolic BP, DBP; Diastolic BP, BMI; Body Mass Index, CI; Confidence Interval, OR; Odds Ratio, AOR; Adjusted Odds Ratio

### Factors associated with non-dipping BP in logistic regression

All factors in univariate analysis demonstrated significant association with non-dipping BP except for sex. At multivariate analysis, age, day SBP, day DBP and night DBP demonstrated a significant association with non-dipping BP. A year increase in age increased the odds of having non-dipping BP by 1.15, p=0.002. A percentage increase in day SBP and night DBP load levels increased the chance of having non-dipping BP by 1.28 and 1.10, respectively, p<0.05. A percentage increase in day DBP reduced the odds of having non-dipping BP by 23% p-value 0.004.

Sex, hypertension status, urine sodium excretion, spot urine sodium, waist circumference and night SBP load level were not significantly associated with non-dipping BP at multivariate analysis. Sex was not significantly associated with non-dipping BP however, compared to males, females had a 48% reduced chance of having non-dipping BP, Table 4.

### Logistic regression analysis of factors associated with non-dipping blood pressure among normotensive individuals

In the normotensive group, we found that participants age, waist circumference and night DBP were the only factors that were significantly associated with non-dipping BP at univariate analysis. However, at multivariable analysis, only age was significantly associated with non-dipping BP, Table 5. A year increase in age increased the odds of having a non-dipping BP among the normotensive participants by 1.16, p=0.02.

**Table 5.** Factors associated with non-dipping BP in normotensive group.

| Variables | OR (95%CI) | P value | AOR (95%CI) | P value |
| --- | --- | --- | --- | --- |
| **Age, years** | 1.20 (1.06,1.35) | **0.002** | 1.16 (1.02,1.31) | **0.02** |
| **Sex** |  | 0.06 |  | 0.47 |
| *Females* | 0.20 (0.03,1.12) |  | 0.34 (0.01,6.54) |  |
| **Waist Circumference, cm** | 1.81 (1.05,3.11) | **0.03** | 2.12 (0.73,6.09) | 0.16 |
| **Night DBP Load Level, %** | 1.05 (1.01,1.09) | **0.01** | 1.04 (0.99,1.09) | 0.11 |

BP; Blood Pressure, DBP; Diastolic BP, CI; Confidence Interval, OR; Odds Ratio, AOR; Adjusted Odds Ratio

**Table 6.** Factors associated with non-dipping BP in hypertension group

| **Variables** | **OR (95%CI)** | **P value** | **AOR (95%CI)** | **P value** |
| --- | --- | --- | --- | --- |
| **Age, years** | 1.13 (1.05,1.23) | **0.002** | 1.21 (0.70,2.09) | 0.47 |
| **Sex** |  |  |  |  |
| *females* | 0.78 (0.24,2.51) | 0.68 | 0.00 (0.00,535880.3)* | 0.23 |
| **BMI, kg/m<sup>2</sup>** | 0.78 (0.62,0.99) | **0.04** | 0.16 (0.01,2.52) | 0.19 |
| **Spot Urine Sodium, mmol/l** | 1.12 (1.03,1.22) | **0.004** | 5.72 (0.33,97.83) | 0.22 |
| **Urine Sodium Excretion, mmol/day** | 1.04 (1.01,1.07) | **0.004** | 0.94 (0.81,1.10) | 0.48 |
| **Day SBP Load Level, %** | 1.06 (1.01,1.11) | **0.01** | 1.37 (0.85,2.20) | 0.18 |
| **Day DBP Load Level, %** | 1.03 (1.00,1.07) | **0.02** | 0.46 (0.16,1.33) | 0.15 |
| **Night SBP Load Level, %** | 1.03 (1.01,1.06) | **0.003** | 1.96 (0.56,6.79) | 0.28 |
| **Night DBP Load Level, %** | 1.03 (1.01,1.05) | **0.002** | 0.92 (0.63,1.42) | 0.73 |

BP; Blood Pressure, SBP; Systolic BP, DBP; Diastolic BP, BMI; Body Mass Index, CI; Confidence Interval, OR; Odds Ratio, AOR; Adjusted Odds Ratio; \*wide confidence interval and AOR 0 due to low sample size at subgroup analysis

### Logistic regression analysis of factors associated with non-dipping blood pressure among hypertensive participants

In the hypertension group, age, spot urine sodium, urine sodium excretion, BMI, Day/Night SBP and Day/Night DBP were significantly associated with non-dipping BP at univariate analysis whereas sex showed no significant association. However, at multivariable analysis, none of the variables were significantly associated with non-dipping BP with all p-values >0.05.

## DISCUSSION

This study aimed to determine the clinical correlates of nocturnal non-dipping blood pressure in persons with and without hypertension. The overall prevalence of nocturnal non-dipping BP was 38.8%. Among those diagnosed with hypertension, non-dipping blood pressure was observed in 54.7% of the participants, while in the normotensive group, 20% exhibited non-dipping blood pressure patterns. These findings mirror a meta-analysis that was conducted in Spain by de la Sierra *et al.* where non-dipping BP was more prevalent in hypertensives in comparison to normotensives [20]. The findings are also similar to a study that was conducted by Ingabire in SSA, where non-dipping BP was notably prevalent among normotensives and hypertensive individuals [13]. Akhtar *et al* study in Qatar revealed a prevalence of non-dipping BP was more prevalent in hypertensives at 74.8% and normotensives at 28.2% [4].

Age was associated with non-dipping BP in this study. This underscores the potential influence of aging on the prevalence of non-dipping patterns, highlighting a need for tailored interventions in older individuals with hypertension. These findings align with studies conducted in Switzerland and Sub-Saharan Africa (SSA) which similarly demonstrated a statistically significant association between non-dipping blood pressure and advancing age, where older participants had increased odds of having non-dipping BP [10,18]. This finding on the age-related influence on non-dipping blood pressure, emphasizes the importance of considering age-specific factors in the management of hypertension. Healthcare providers should incorporate age-related changes in blood pressure regulation into their decision-making processes, ensuring tailored interventions for older individuals.

The study’s most notable associations are observed with daytime and nighttime blood pressure load levels. Daytime systolic blood pressure (SBP) load level indicates 28% increased odds of non-dipping patterns, emphasizing the role of heightened daytime SBP load in the occurrence of non-dipping. The research findings align closely with those of a study conducted in Brazil, a country characterized by low to medium income levels. Similarly, Zambia, identified as a low-income country, also reflects comparable outcomes. A similar study conducted in this context revealed a statistically significant association between non-dipping blood pressure and daytime systolic blood pressure (SBP) load among non-dippers and reverse dippers. This consistency across different socioeconomic settings underscores the robustness and generalizability of the observed relationship between non-dipping blood pressure and daytime SBP load, regardless of the economic context [23]. The findings show further consistency with study that were done in high income countries, Spain and Italy, whose findings revealed an association of daytime systolic blood pressure (SBP) load level to development of non-dipping BP [21,22].

The study’s findings carry substantial public health and clinical implications. The observed high prevalence of non-dipping blood pressure, particularly among individuals with hypertension, underscores the imperative for comprehensive monitoring of nocturnal blood pressure patterns. This heightened prevalence emphasizes the need for vigilant cardiovascular risk assessment and individualized treatment strategies in hypertensive patients, acknowledging the potential impact of non-dipping blood pressure on long-term outcomes. Public health interventions should focus on raising awareness about the significance of nocturnal blood pressure monitoring, facilitating early detection of non-dipping patterns, and tailoring interventions to mitigate associated cardiovascular risks.

The associations with daytime blood pressure load, both systolic and diastolic, underscore the critical role of daytime blood pressure control in preventing nocturnal hypertension. This highlights the need for aggressive management of hypertension during waking hours, not only to address daytime cardiovascular risks but also to potentially mitigate the development of non-dipping patterns and associated adverse events during sleep.

In contrast, daytime diastolic blood pressure (DBP) load correlated negatively with non-dipping blood pressure indicating a potential protective effect of elevated daytime DBP load against non-dipping. This finding is not consistent with any other findings and beyond the scope of our study to ascertain the underlying reason. Nighttime DBP load was positively correlated with non-dipping blood pressure. These results are similar to previous studies, where it was observed that individuals characterized as non-dippers exhibited a notably higher average 24-hour systolic and diastolic blood pressure [26–28].

### Study limitations and strengths

The study did not consider the dietary habits and hydration state or time for the collection of urine for sodium excretion. Various confounding factors could have influenced the outcome of interest including, dietary habits. Different foods have different sodium, which can affect blood pressure. Also, the volume and timing of fluid intake could influence urine production and sodium excretion. The study further lacked longitudinal data. It employed a cross-sectional study design, while useful it presents certain limitations. A cross-sectional design, by capturing only a single point in time, might miss essential fluctuations blood pressure and sodium excretion, and their potential impact on the relationships of study. Another important limitation is the small sample size especially affecting subgroup analysis of the hypertension and normotensive groups. This can be seen from the wide confidence interval of the sex variable in the multivariable analysis. These data should therefore be interpreted with caution and will need to be validated by a large sample size. Recruiting participants was especially hard due to the requirements for ambulatory blood pressure monitoring requiring participants to wear the blood pressure monitoring device for 24 hours.

The study demonstrated notable strengths, including its comprehensive data collection covering sociodemographic and clinical variables such as age, sex, hypertension status, lifestyle factors, and urine sodium excretion, providing a thorough assessment of potential correlates of non-dipping blood pressure. Furthermore, the application of multivariate logistic regression enabled the simultaneous exploration of multiple factors associated with non-dipping blood pressure while controlling for potential confounders, bolstering the reliability of the findings. Its clinical relevance is underscored by its investigation into the correlates of non-dipping blood pressure, a critical aspect for hypertension management and cardiovascular risk assessment. By focusing on non-dipping blood pressure, a crucial prognostic indicator for cardiovascular health, the study contributes valuable insights into its prevalence and associated factors among individuals with and without hypertension.

## Conclusion

The prevalence of non-dipping blood pressure was high in the study and especially among persons with hypertension. This study provides insights into the intricate links between blood pressure pattern, sociodemographic and clinical characteristics. However, there is a need for further studies to comprehensively understand the mechanisms behind non-dipping BP and its associated characteristics.

## Declarations

### Ethics approval and consent to participate

Ethical clearance for the research was secured from the Mulungushi University School of Medicine and Health Sciences-Research Ethics Committee (MUSoMHS-REC) (Assurance No. FWA: 0002888 IRB: 00012281 of IORG0010344) on 27^th^ January, 2023. Official approval to carry out the study was granted by the National Health Research Authority (NHRA). At no time over the course of this study was it possible to determine the identity of any participant included in the database. There was no breach of confidentiality to be reported to the Mulungushi University School of Medicine and Health Sciences Research Ethics Committee during the course of the study.

### Consent for publication

Not applicable

### Availability of data and materials

All data produced or examined in the course of this study have been incorporated into this published article. Requests for additional data can be made to the corresponding author.

### Author’s contributions

SL and SKM conceived the study. BMH, OLMM, SDZ and SKM contributed to the writing of the manuscript. SL is the principal investigator. SKM is the senior author. All authors read, provided feedback, and approved the final manuscript.

## Data Availability

All data produced are available online at

https://www.editorialmanager.com/pone/download.aspx?id=35566510&guid=83693d2f-fb5f-49c2-8728-8d32b3d8c1a9&scheme=1

## Acknowledgments

We are very grateful to all Laboratory personnel and the Medical Superintendent’s office at Chikankata Mission General Hospital for their support and enthusiasm for research projects. We would also like to thank the CMGH hypertension clinic for their continued support and assistance during the data collection process.

## Abbreviations

ABPM: Ambulatory Blood Pressure
ABP: Ambulatory Blood Pressure Monitoring
ACC: American College of Cardiology
AHA: American Heart Association
ACE: Angiotensin-Converting Enzyme
ANT: Autonomic Nervous System
DBP: Diastolic Blood Pressure
BP: Blood Pressure
CHF: Congestive Heart Failure
CMGH: Chikankata Mission General Hospital
CVD: Cardiovascular Diseases
HDL-C: High Density Lipoprotein-Cholesterol
INTERSALT: International Study of Salt and Blood Pressure
IRB: Institutional Review Board
NDBP: Non-Dipping Blood Pressure
NHRA: National Health Research Authority
RAAS: Renin-Angiotensin-Aldosterone System
SBP: Systolic Blood Pressure
SSA: Sub-Saharan Africa
WC: Waist Circumference

